# A Systems Approach Identifies Key Regulators of HPV-Positive Cervical Cancer

**DOI:** 10.1101/2020.05.12.20099424

**Authors:** Musalula Sinkala, Mildred Zulu, Panji Nkhoma, Doris Kafita, Ephraim Zulu, Rabecca Tembo, Zifa Ngwira, Victor Daka, Sody Munsaka

**Author notes:** Contributed equally (Musalula Sinkala, Mildred Zulu, Panji Nkhoma).

## Abstract

Cervical cancer has remained the most prevalent and lethal malignancy among women worldwide and accounted for over 250,000 deaths in 2019. Nearly ninety-five per cent of cervical cancer cases are associated with persistent infection with high-risk Human Papillomavirus (HPV), and seventy per cent of these are associated with viral integration in the host genome. HPV-infection imparts specific changes in the regulatory network of infected cancer cells that are of diagnostic, prognostic and importance. Here, we conducted a systems-level analysis of the regulatory network changes, and the associated regulatory proteins thereof, in HPV-positive cervical cancer. We applied functional pathway analysis to show that HPV-positive cancers are characterised by perturbations of numerous cellular processes, predominantly in those linked to the cell cycle, mitosis, cytokine and immune cell signalling. Using computational predictions, we revealed that HPV-positive cervical cancers are regulated by transcription factors including, SOX2, E2F, NANOG, OCT4, and MYC, which control various processes such as the renewal of cancer stem cells, and the proliferation and differentiation of tumour cells. Through the analysis of upstream regulatory kinases, we identified the mitogen-activated protein kinases; among others, MAPK1, MAPK3 and MAPK8, and the cyclin-dependent kinases; among others, CDK1, CDK2 and CDK4, as the key kinases that control the biological processes in HPV-positive cervical cancers. Taken together, we uncover a landscape of the key regulatory pathways and proteins in HPV-positive cervical cancers, all of which may provide attractive drug targets for future therapeutics.

## Introduction

Cervical cancer affects more than 500,000 women and causes more than 300,000 deaths per year in women worldwide [1,2]. Nearly ninety per cent of cervical cancer cases occur in less developed countries, where the burden of disease is disproportionately high. The disease ranks as the number one cause of female cancer mortality accounting for about a tenth of all deaths in women due to cancer [1,2].

Cervical cancer is associated with persistent infection of high-risk human papillomaviruses (HPV). Identifying cervical cancer as having a viral aetiology has substantive consequences both for its treatment and for its prevention [1]. It is anticipated that over ninety-five per cent of cervical cancer cases will be prevented in the future by eliminating persistent infections with more than 15 oncogenic or high-risk genotypes of HPV. However, currently, cervical cancer remains mostly incurable [3]. As with other cancer-associated viruses, persistent infection of high-risk HPV genotypes is necessary for carcinogenesis of cervical cancer [4–6]. Recently, efforts have revealed that HPV virus oncogenes inactivate the tumour suppressor proteins p53 and pRB, leading to increased genomic instability, and among other cancer hallmarks, and in some cases, integration of HPV into the host genome [6,7]. The molecular characterisation of cervical cancer by the Cancer Genome Atlas project (TCGA) and others, has provided us with a multi-dimensional understanding of the transcriptomic, genomic, proteomic, and epigenetic landscape of the disease [8–13]. These recent efforts have led to, for example, the identification of transcriptional changes of the molecularly distinct disease subtypes, the mutational landscape of cervical cancer [4,9,10], and the drug response and specific genetic vulnerabilities of the cervical cancer cells [14].

Despite progress made, we have barely begun to understand how some of these alterations could be leveraged to achieve effective therapeutics, owing to that we lack a multiscale understanding of how the regulatory networks of these cancers are rewired in the presence of HPV-infection. Furthermore, we are yet to identify the dysregulated pathways and the key regulators thereof that are of clinical relevance for HPV-associated cervical cancers. Here, therefore, we present a systems-level analysis of invasive cervical cancer with a focus on identifying novel alterations in the regulatory network of HPV-positive cervical cancer, the key transcription factors and kinases that may drive tumorigenesis, and thus which may all serve as markers for future therapeutic strategies.

## Results

### Clinical characteristics HPV-positive cervical cancer

We assembled a TCGA cervical cancer dataset comprising clinical information for 180 patients together with their associated mRNA transcription data. We segregated the cervical cancer samples into two groups; those that were reported HPV-positive (156 samples) and those that were HPV-negative (24 samples). We found no differences in disease outcome after the first course of treatment between HPV-positive and HPV negative cervical cancer patients (Figure1a, also see Figure S1a). Here, we also found that the reported vital statistics were not statistically different. Eighty-one per cent and eighty-three per cent of the patients were alive at the end of the follow-up period, for the patients who had HPV-positive and HPV-negative cervical cancer, respectively (Figure 1b).

**Figure 1:**
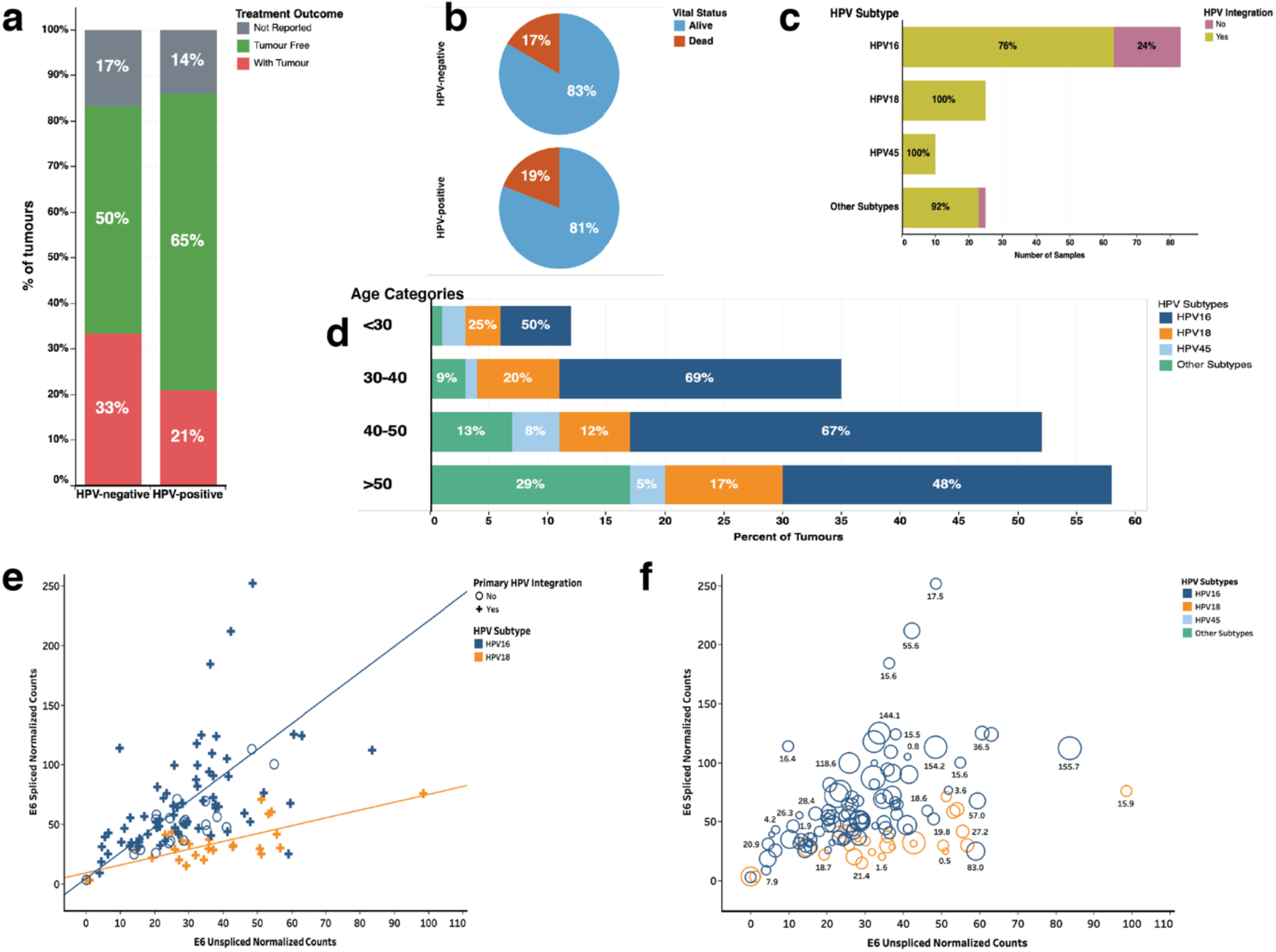
**(a)** Comparison of the clinical outcomes after the first course of treatment across the HPV-positive and HPV-negative cancer patients. **(b)** Pie chart showing the vital statistics after the first course of treatment across the HPV-positive and HPV-negative cancer patients. (c) Frequency of HPV subtypes in the cancer samples. The colours show the split for samples with HPV viral DNA that is integrated into the host genome. **(d)** Distribution of HPV-subtypes of different aged categories of tumours of patients who have HPV-positive cervical cancer. **(e)** Pearson’s linear correlation between the normalised E6 spliced mRNA transcripts and the normalised E6 unspliced mRNA transcripts measured in HPV-positive cancers. The regression lines are plotted separately for HPV-subtypes 18 and 16. The marker shape shows labelled based on the primary HPV viral DNA integration into the host genome. **(f)** E6 unspliced normalised Counts E6 versus E6 spliced normalised counts. The colour shows details about HPV-subtype. The marks size shows details of the duration of overall survival (in months) of the afflicted patients. The marks are labelled with the duration of the overall survival.

The number of HPV subtypes included HPV-16 (58% of the samples), HPV-18 (18%), HPV-45 (7%), other subtypes (< 17%) (Figure S1a). HPV-45 and HPV-18 showed the highest level of integration into the host genome (100% for both virus subtypes), followed by other HPV-subtypes (92% integration) and the least level of integration into the host genome was seen among the HPV-16 subtype (76% integration) (Figure 1c). We found that 29% of cervical cancer patients older than 50 years had tumours that were associated with other HPV subtypes compared to the other age categories (< 30 years, 30 to 40 years and 40 to 50 years; 7% of samples), with a statistically significant difference, χ2 = 4.0, p = 0.045 (Figure 1d). We found no other statistically significant association between the HPV subtypes and the different age categories of cervical cancer patients.

We assessed the relationship between E6-spliced and E6-unspliced mRNA transcripts in the cervical cancer samples to find a significant positive correlation (r = 0.16, p < 0.0001; Figure 1e). Here, we found that the Pearson linear correlation coefficient for E6-spliced verse E6-unspliced was higher for tumours infected with the HPV-16 subtype (r = 0.59, p < 0.0001) compared to tumours infected with HPV-18 subtype (r = 0.29, p < 0.0001). Furthermore, we found no significant association between the levels of E6 spliced and unspliced mRNA transcript and the survival outcomes of the patients who had cervical cancer (Figure 1f and Figure S1b). All the clinical information is presented in supplementary file 1.

### Up-regulated genes in HPV-positive cervical cancer

We used the moderated t-test and the empirical Bayes methods to identify the differentially expressed genes in either HPV-positive or HPV-negative cervical cancers compared to the normal, non-cancerous samples. Here, we found 2,033 mRNA transcripts up-regulated in HPV-positive tumours. Among the most significantly upregulated genes were PTTG3P (log 2-fold-change [log2FC] = 11.7, q < 1 × 10^−50^), MMP12 (Log2FC = 10.6, q < 1 × 10^−50^), and BCAR4 (Log2FC = 9.5, q < 1 × 10^−50^; see Supplementary File 2 for all the differentially expressed genes). Most of the genes that we found significantly up-regulated in HPV-positive tumours have well-defined roles that are linked to neoplastic processes and response to viral infections. For instance, the overexpression of the pseudogene PTTG3P (Putative pituitary tumour-transforming gene 3-protein) is linked to carcinogenesis, tumour progression and metastasis in epithelial tissues in most malignancies including those of the cervix, oesophagus, breast, liver, bladder and lung [15–18]. MMP12 encodes the Macrophage metalloelastase which has antiviral immune activity and involvement in the malignant transformation of among others, cells of the cervix and skin [19–22].

### Regulatory pathways and cellular processes of HPV-positive cervical cancer

We first set out to identify the dysregulated Reactome pathways among the cervical cancer samples by using the significantly up-regulated mRNA transcripts (q-value < 0.05 and Log2FC > 2). Here, to identify Reactome pathway enrichments in cervical tumours, we used three lists of up-regulated mRNA transcripts: 1) only those transcripts up-regulated in HPV-positive tumours, 2) transcripts up-regulated in both HPV-positive and HPV-negative tumours, and 3) transcripts up-regulated only in HPV-negative tumours (Figure 2).

**Figure 2:**
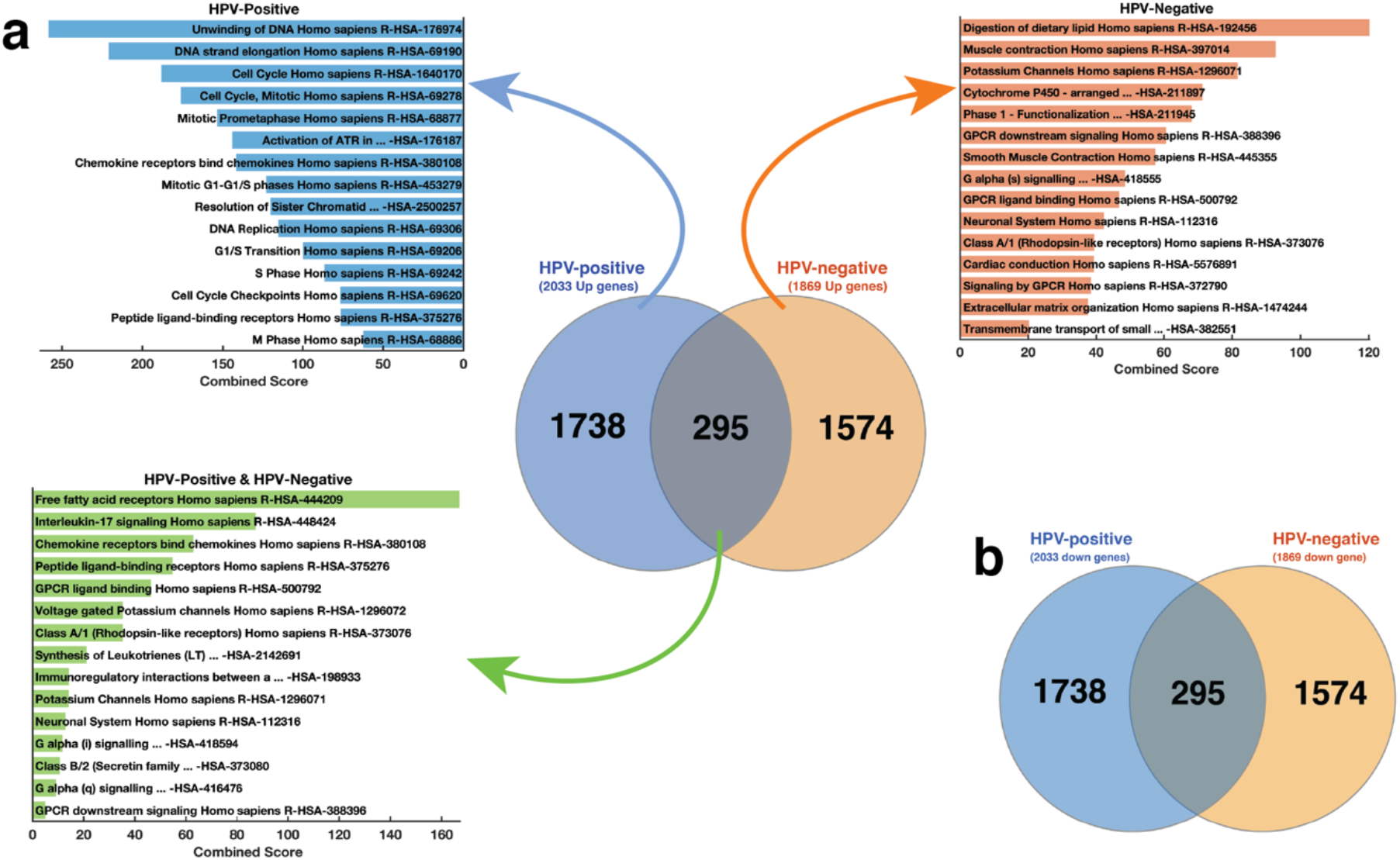
**(a)** Gene enrichment analysis plots. Top left; the top-15 Reactome pathways enriched for in HPV-positive samples. The analysis is based on the 1,738 mRNA transcripts that are up-regulated only in these samples. Centre; The distribution of significantly up-regulated mRNA transcripts among HPV-positive and HPV-negative samples. Top right; the top-15 Reactome pathways enriched for in HPV-negative samples. The analysis is based on the 1,864 mRNA transcripts that are up-regulated only in these samples; Bottom left; the top-15 Reactome pathways enriched for in both the HPV-positive and HPV-negative cervical cancers. Refer to supplementary file 3 for the complete list of Reactome pathways that represent significantly enriched genetic alterations in the HPV-positive, HPV-negative, both cervical cancers. **(b)** The distribution of significantly down-regulated mRNA transcripts among HPV-positive and HPV-negative samples.

Using the 1,738 mRNA transcripts that we found up-regulated only in HPV-positive cervical cancers, we found that HPV-positive cancers exhibited a significant enrichment for 121 unique Reactome pathways (Supplementary File 3). Among the top-ranked Reactome pathways where those associated with the Cell Cycle processes (Combined Score [CS] = 188.4, q = 1.05 × 10^−26^), Mitotic G1-G1/S phases (CS = 122.7, 1.84 × 10^−12^), and DNA Replication (CS = 115.4, p = 4.33 × 10^−11^; Figure 2a). Conversely and unexpectedly, by focusing on the 1,574 mRNA transcripts that we found up-regulated only in HPV-negative cancers, we revealed that HPV-negative cancers exhibit significant enrichment for only 31 Reactome pathways. Here, most of the pathways enriched for are associated with G-protein coupled receptor signalling (Figure 2a).

Furthermore, we focused on the 295 mRNA transcripts that were up-regulated in both HPV-positive and HPV-negative tumours to reveal significant enrichment for only three Reactome pathways. These were associated with G-protein coupled ligand binding (CS = 46.4, q = 0.001), Peptide ligand-binding receptors (CS = 54.8, q = 0.005), and Rhodopsin-like receptors activity (CS = 35.4, q = 0.014; see Supplementary File 3).

Surprisingly, we found that all the 2,010 mRNA transcripts that were significantly down-regulated in HPV-positive samples were also significantly down-regulated in HPV-negative samples (Figure 2b). An additional 23 mRNA transcripts were down-regulated in the HPV-negative tumours. Here, our results suggest that while the pathways whose activities are increased are distinctive among the cervical tumours that are HPV-positive and HPV-negative, those with reduced activity are comparable for both.

For the remaining analyses, we used the 1,738-mRNA transcript that we found up-regulated only in HPV-positive cervical tumours. Using these, first, we sought to identify the KEGG pathways, Gene Ontology (GO) terms Biology Processes and GO-term Molecular Function that are enriched for in HPV-positive cervical tumours. Here, our analyses revealed that the HPV-positive cancers are significantly enriched for 23 KEGG pathways, 175 GO-term Biological Processes and 25 GO-term Molecular Functions (Table 1, Supplementary File 3).

**Table 1:** Regulatory pathways and functional processes in HPV-positive cervical cancer

| PATHWAY OR FUNCTIONAL PROCESS | P-VALUE |
| --- | --- |
| KEGG Pathways |  |
| Cell cycle | $8.3 \times 10^{-12}$ |
| Cytokine-cytokine receptor interaction | $2.4 \times 10^{-10}$ |
| DNA replication | $8.5 \times 10^{-10}$ |
| Cell adhesion molecules (CAMs) | $5.9 \times 10^{-05}$ |
| Hematopoietic cell lineage | $1.4 \times 10^{-04}$ |
| GO Biological Processes |  |
| Cytokine-mediated signalling pathway (GO:0019221) | $9.3 \times 10^{-16}$ |
| Chemokine-mediated signalling pathway (GO:0070098) | $1.0 \times 10^{-11}$ |
| The inflammatory response (GO:0006954) | $1.3 \times 10^{-11}$ |
| DNA replication (GO:0006260) | $1.7 \times 10^{-11}$ |
| Lymphocyte chemotaxis (GO:0048247) | $1.2 \times 10^{-10}$ |
| GO Molecular Function |  |
| Chemokine activity (GO:0008009) | $4.1 \times 10^{-12}$ |
| Chemokine receptor binding (GO:0042379) | $2.0 \times 10^{-11}$ |
| Cytokine activity (GO:0005125) | $2.3 \times 10^{-10}$ |
| UDP-glycosyltransferase activity (GO:0008194) | $9.3 \times 10^{-07}$ |
| CXCR chemokine receptor binding (GO:0045236) | $3.5 \times 10^{-06}$ |

Among the KEGG pathways, the most significantly enriched for were those associated with the Cell cycle (p = 8.33 × 10^−12^), Cytokine-cytokine receptor interaction (p = 2.39 × 10^−10^) and DNA replication (p = 8.46 × 10^−10^). For the GO-term Biological Processes, those that we found most significantly enriched for in HPV-positive tumours include the Cytokine-mediated signalling pathway (p = 9.34 x 10-16), Chemokine-mediated signalling pathway (p = 1.02 × 10^−11^), and Inflammatory response (p = 1.29 × 10^−11^). Furthermore, for HPV-positive tumours, the top-three significantly enriched GO-term Molecular Function were those linked to Chemokine activity (p = 4.08 × 10^−12^), chemokine receptor binding (p = 2.0 × 10^−11^), and cytokine activity (p = 2.29 × 10^−10^).

We connected the significantly enriched KEGG pathway terms, GO-term Biological Processes, and GO-term Molecular Functions using known gene interactions (see Methods Sections) to reveal the overall enrichment landscape of HPV-positive cervical cancer (Figure 3). Here, we found that HPV-positive tumours are primarily driven by cytokine signalling and cell cycle processes.

**Figure 3:**
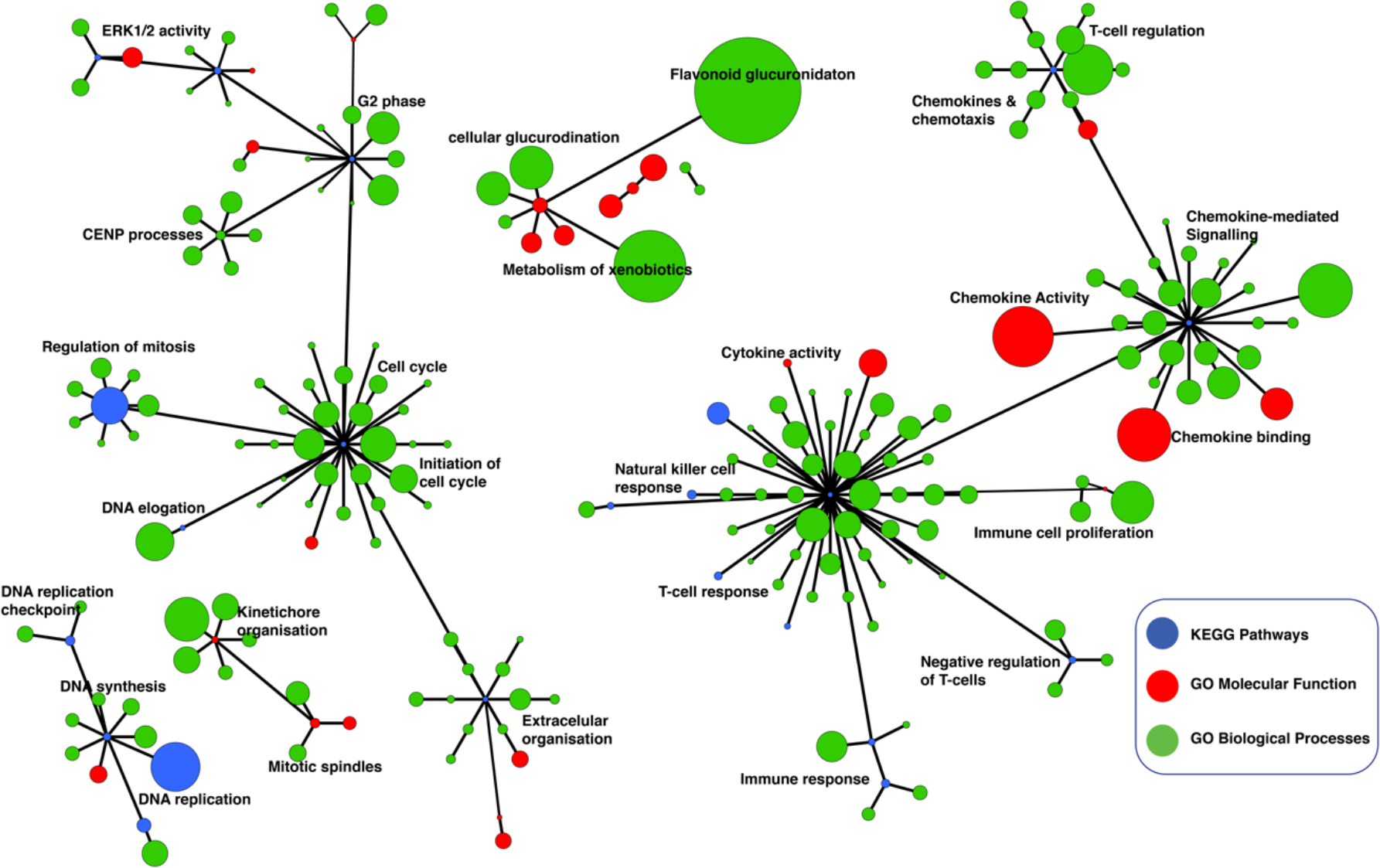
Network of Gene Ontology (GO) terms Molecular Functions, GO-term Biological Processes and KEGG pathways found enriched for in HPV-positive cervical cancers. Enrichr was used to obtain enriched GO-terms and KEGG pathways. The three enrichment results were combined to create an integrated network enriched functional process and pathways that were visualised in yEd (refer to the Methods section). Each node represents a GO-term Biological Process (blue), GO-term Molecular Function (red), and KEGG pathway (green). The nodes that represent similar function processes are clustered together and connected by edges. The thickness of the edges represents the number of shared genes between the nodes. The size of each node denotes the gene set size of the represented GO-term or KEGG pathway. Refer to supplementary file 3 for the complete list of KEGG pathways and GO-terms that represent significantly enriched regulatory networks in the HPV-positive cervical cancers.

Overall, these findings uncover the distinct molecular mechanisms and signalling pathway perturbations which mediate oncogenic behaviour of cervical tumours that are associated with HPV infection.

### Transcription factors that regulate HPV-positive cervical cancer

We used the complete list of the mRNA transcripts that we found significantly up-regulated in HPV-positive cervical cancer samples compared to the normal cervical tissues to compute enrichment for transcription factors that likely regulate the observed differential gene expression pattern. Using the *in silico* approach, Chromatin Immunoprecipitation Enrichment Analysis (ChEA) [23], we predicted 35 significant (p < 0.05) transcription factors that likely regulate the molecular processes that govern HPV-positive cervical cancer (Figure 4a; also see Supplementary File 4). Here, the top-ranking transcription factors were SOX2 (CS = 92.2, p = 1.51 × 10^−24^), E2F4 (CS = 73.3, p = 9.24 × 10^−13^), E2F1 (CS = 44.6, p = 1.21 × 10^−10^) and POU5F1 (CS = 35.0, p = 2.82 × 10^−08^).

**Figure 4:**
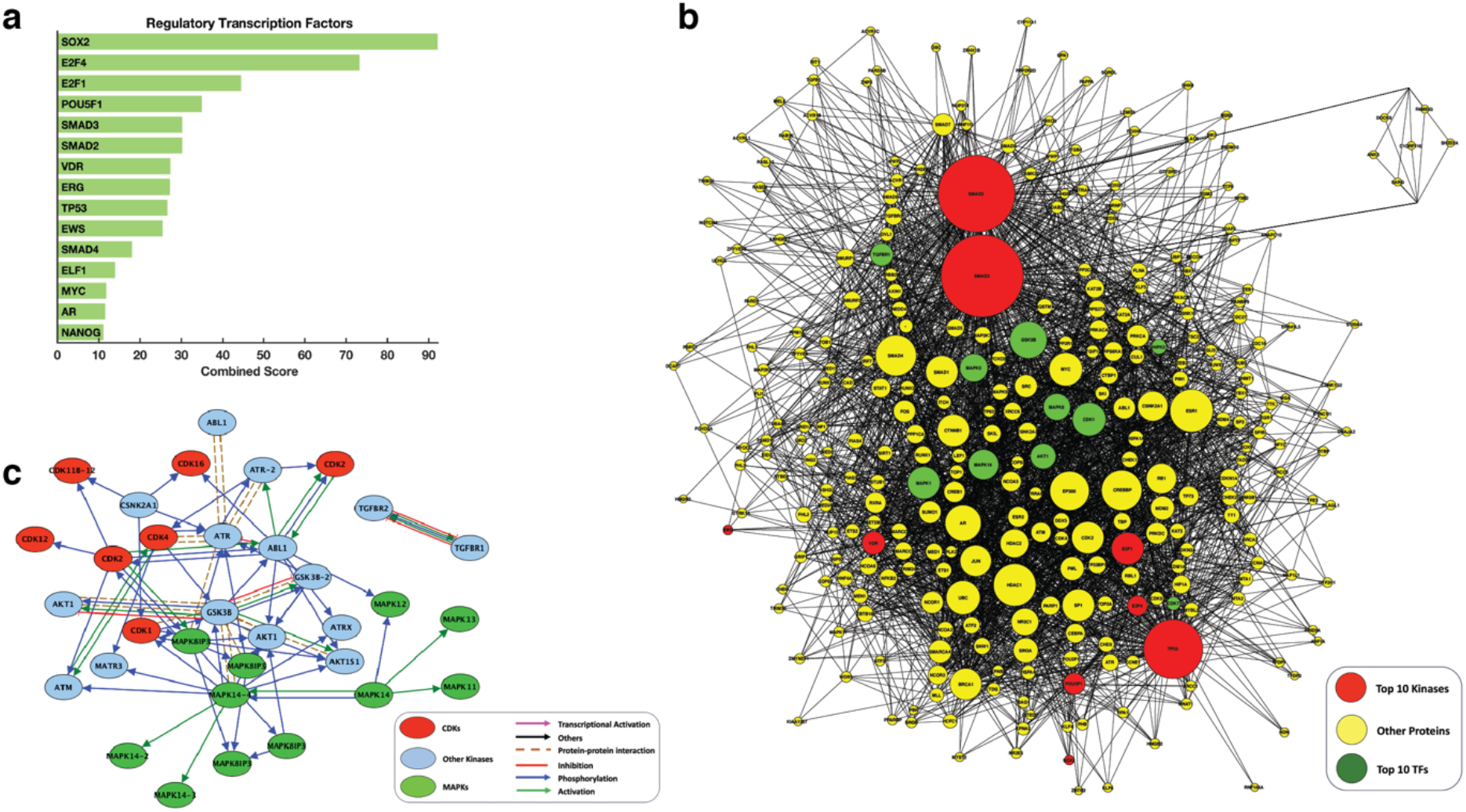
**(a)** the top-15 predicted regulatory transcription factors. **(b)** Intermediate protein-protein interaction subnetwork connecting transcription factors to the upstream regulatory proteins: the sub-network has 180 nodes with 1816 edges. The network is enriched for co-regulators, kinases and transcription factors that are experimentally verified to interact physically. **(c)** Connectivity of the top-twenty predicted kinases. Red nodes represent the cyclin-dependent kinases, the green node represent the mitogen-activated protein kinase, and cyan nodes represent other regulatory kinases. Blue nodes kinases other than the cyclin-dependent kinases or mitogen-activated protein kinases.

Several studies show that SOX2 is overexpressed in cervical tumours and is associated with cervical neoplasia [24–26]. POU5F1 binds to the octamer motif (5’-ATTTGCAT-3’) and controls the expression of several genes involved in embryonic development, oncogenesis, tumour metastasis and disease aggressiveness in several human cancers [27–29]. However, little is known about the specific roles of POU5F1 in cervical cancer. Conversely, the interplay between E6/E7 and E2F1/E2F4 in modulating the mitotic pathway, p53 pathway, pRB pathway and among other pathways is well documented [30–34].

### Regulatory kinases of HPV associated cervical cancer

We leveraged prior-knowledge protein interactions to connect the transcription factors identified using the ChEA analysis to upstream interactors and regulators to yield a protein-protein interaction sub-network (Figure 4b). The sub-network has 309 nodes that are connected by 3,299 edges. This sub-network is enriched for co-regulators, kinases and interactors of the transcription factors that drive the gene expression signature of cervical cancers infected with HPV.

Next, we used Kinase Enrichment Analysis (KEA) [35] to link the proteins in the protein-protein interaction sub-network to the protein kinases that likely phosphorylate them. Here, our result was a ranked list of protein kinases that likely regulate the transcriptome signature of HPV-positive cervical tumours. Among the top-ten ranked kinases were four mitogen-activated protein kinases (MAPKs):

MAPK1 (p = 4.82 × 10^−15^), MAPK3 (p = 6.56 × 10^−14^), MAPK8 (p = 1.71 × 10^−10^), and MAPK14 (p = 2.68 × 10^−07^; Figure 4c and Figure 5, also see Supplementary File 4). The MAPK pathway genes are altered in cervical cancers, and among other human cancers in which the MAPKs are the chief signal transduction proteins that regulate oncogenic behaviour such as cell proliferation, cell differentiation, cell survival, cancer metastasis, and resistance to drug therapy [36–40]. Accordingly, we suggest that the MAPK signalling pathways may present inflexion points for targeted therapies aimed at successfully treating HPV associated cervical cancer.

**Figure 5:**
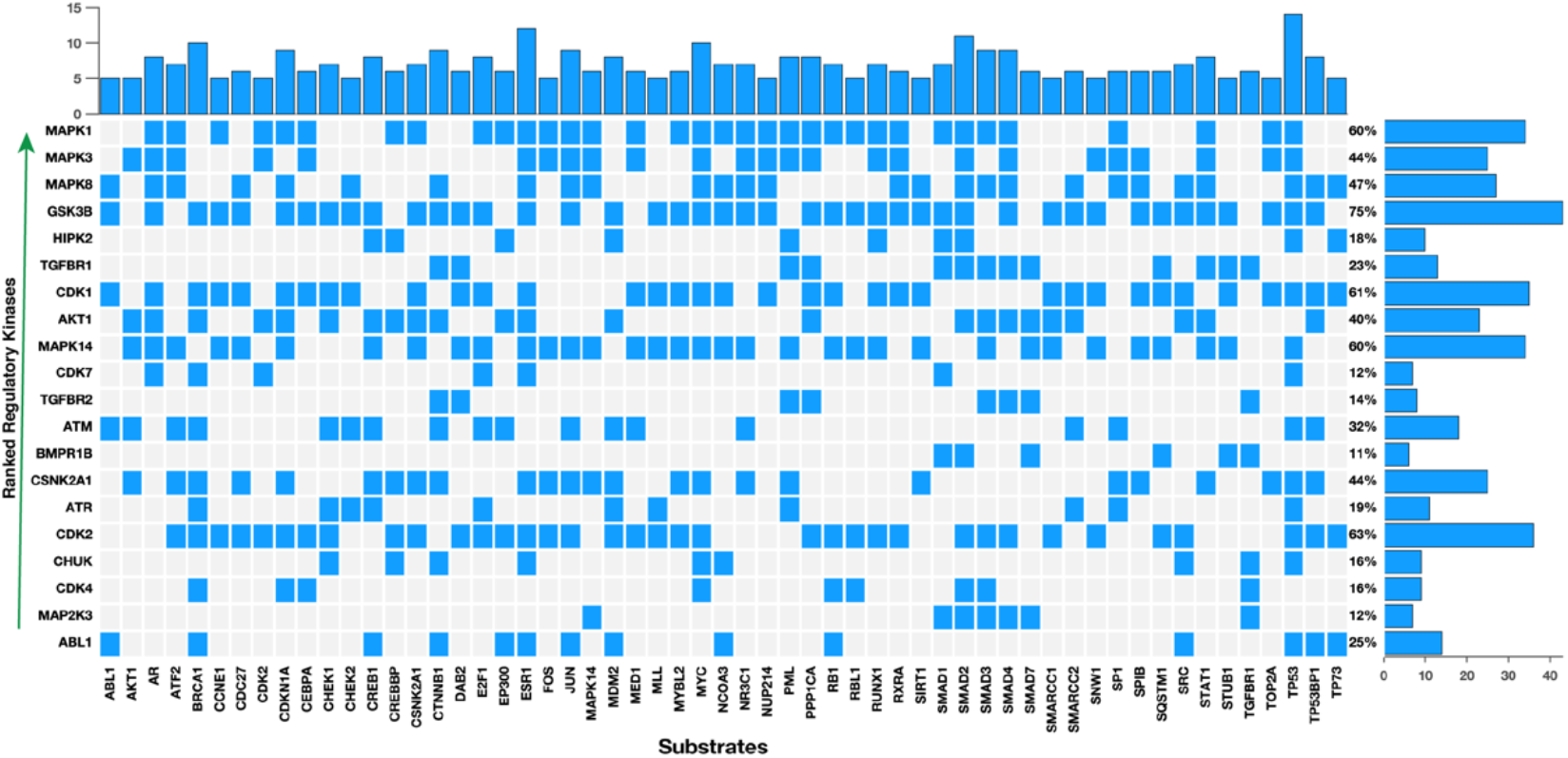
the top-ten predicted regulatory kinases ranked according to their combined statistical score based on the number of substrates they phosphorylate within a protein-protein interaction subnetwork. Shown along the columns of the plot are proteins which are the substrates for kinases along the rows.

## Discussion

We conducted a systems-level analysis to show that the biological underpinnings of HPV-positive cervical cancers are different from those of HPV-negative cancers. Our analysis revealed 121 significantly dysregulated Reactome pathways in HPV-positive cervical cancers and 31 pathways in HPV-negative cervical cancers. This finding suggests that infection with high-risk HPV genotypes additionally dysregulates several other signalling pathways, some of which may enable cancer cells to either acquire resistance or become more responsive to therapies [41–44]. We identified that the most prominent cell signalling aberrations in the HPV-positive cervical cancers were within the cell cycle and mitosis associated pathways (Figure 2a); hinting that these cancers might respond to drugs that target the cell cycle. As expected, cervical cancers have been shown to exhibit a high therapeutic response to drugs that arrest cell cycle processes [14,45]. In this regard, we show that although alterations of multiple cancer hallmarks are required for cervical oncogenesis, those related to cell cycle processes predominantly regulate the oncogenesis in HPV-positive cervical cancer [46].

We showed that the clinical characteristics of the patients are vastly comparable for both the patients afflicted with either HPV-positive or HPV-negative cancer. Consistent with our findings, others have also shown that the overall survival outcomes of cervical cancer patients are likely not influenced by HPV infection [47]. Patients who have HPV-inactive cervical cancer have been shown to have worse outcomes compared to those who have HPV-active cervical cancer [48]. Furthermore, these findings assert the need to identify the molecular variances between cancer subtypes, which are of greater relevance to the chemotherapeutic responses of the cancers [49–53].

We integrated our findings of the altered regulatory signalling pathways (KEGG pathways), biological processes, and molecular functions to reveal the integrated regulated network of HPV-positive cervical cancers (Figure 3). Interestingly, we observed that besides alterations in the Cell cycle-related processes, HPV-positive cervical cancers show pervasive dysregulation of, among others, Cytokine-mediated processes, DNA replication, and Chemokine-mediated signalling pathway. Cytokine dysregulation in cervical cancer is a hallmark of persistent infection with high-risk HPV caused by the immunosuppressive tumour microenvironment mediated by several proinflammatory cytokines such as IL-4, IL-6, TNF-α and IFN-γ [54–57]. Strategies aimed at reversing the cytokine-related treatments are presently either being tested in clinical trials or are already in use as cervical cancer therapies with varied success [58–60].

Our computational predictions identified, among the 35 significant transcription factors, SOX2, NANOG, POU5F(OCT4), and MYC as crucial regulators of HPV-positive cervical cancer. Interestingly, the transcription factors SOX2, NONOG, MYC and OCT4 are stem cell markers that regulate the development of signalling networks including the apoptosis and cell-cycle pathways, and other pathways associated with the phenomenal proliferation and self-renewal of cells [61–63]. Since transcription factors are generally regarded as “undruggable”; we could indirectly restrain the activity of those we have identified here by inhibiting their upstream regulatory kinases [64,65]. Concretely, we have identified several kinases of the MAP-kinase pathways, which control cell cycle-related processes including cellular proliferation, and differentiation [66–68], and kinases that directly regulate the cell cycle such as CDK1, CDK2, CDK4 and CDK7 [69,70]. These kinases, and among others, represent credible targets for small molecule inhibitors, including CDK and MAPK inhibitors, that might be useful for controlling the disease. Notably, the pervasive alterations that we have observed in the cell cycle processes and cytokine-mediated activity provide the rationale for potentially treating HPV-positive cervical cancers using an approach that synchronously targets all these pathways.

Altogether, our analysis revealed the perturbed regulatory signalling pathways, transcription factors and protein kinases that likely govern the behaviour of HPV-positive cervical cancers, many of which are therefore potential targets for anticancer drugs.

## Methods

We obtained uniformly processed mRNA expression data of 25 non-cancerous cervical tissue samples and 294 cervical cancer samples by the TCGA from the ARCHS4 (all RNA-seq and ChIP-seq sample and signature search) database [71,72]. These data included mRNA transcription levels of cervical cancer samples and comprehensive de-identified clinical and sample information. The clinical information of the samples is summarised in supplementary file 1.

### Identification of differentially expressed mRNA transcripts

First, we removed the genes with transcript levels less than 1 Fragment Per Kilobase Million across all samples from mRNA transcription data. Then we applied the moderated student t-test based on the negative binomial model [73,74] to identify differentially expressed mRNA transcripts between 1) HPV-positive cervical tumours and non-cancerous cervical tissues and 2) HPV-negative cervical cancer tumours and non-cancerous cervical tissues (see Supplementary File 2). Furthermore, we defined up-regulated and down-regulated mRNA transcripts for both comparisons as those with the Benjamin-Hochberg corrected p-values [75] less than 0.05 and log2 fold-change either > 2 for the up-regulated transcripts or <-2 for the downregulated transcripts.

### Pathway and functional enrichment analyses

We assessed enrichment for various Reactome pathways [76,77] in cervical cancers by querying Enrichr [78,79] with three gene lists for those: 1) that we found up-regulated only in the HPV-positive tumours, 2) up-regulated only in the HPV-negative tumours and 3) up-regulated in both the HPV-positive and HPV-negative tumours (see Supplementary File 3).

Furthermore, we used the list of genes that we found up-regulated only in HPV-positive cervical tumours to evaluate the KEGG pathways [80], GO-term Molecular Function and GO-term Biological Processes [81] that are enriched for in HPV-positive cervical samples. We used a custom MATLAB script to connect the enriched KEGG pathway terms, GO-term Molecular Function and GO-term Biological Processes into an integrated regulatory enrichment map. Then we used software yEd to visualise the integrated enrichment landscape of the HPV-positive cervical tumours.

### Prediction of the regulatory transcription factors and kinases

We identified the transcription factors and kinases that regulate the oncogenic behaviour of HPV-positive cervical cancer using a computational approach that is implemented in the software Expression2 Kinases (X2K) [82]. X2K employs a network-based reverse engineering approach to make the prediction based on prior-knowledge network data.

Here, we used a list of genes that are significantly up-regulated only in the HPV-positive tumours compared to the HPV-negative tumours. Then we used the Chromatin Immunoprecipitation (ChIP) Enrichment Analysis (ChEA; 2016) [23] module of X2K to predict the upstream regulatory transcription factors that likely lead to observed transcription signature of the HPV-positive tumours (see Supplementary File 4). Next, we constructed a protein-protein interaction subnetwork by connecting the top-10 predicted transcription factors to their upstream regulators. Here, we used prior knowledge interactions from the Biological General Repository for Interaction Datasets [83], Kinase enrichment analysis database [35], KEGG pathways [84] and the Human Interaction database. Finally, we applied Kinase Enrichment Analysis to analyse the protein-protein interaction sub-network for protein kinases which are enriched for based on the number of targets that each kinase phosphorylates within the sub-network (KEA; 2015) [35]. See supplementary file 4 for a full list of the predicted kinases and their rankings based on the combined score.

### Statistical Analyses

All the analyses that we have presented here were performed in MATLAB version 2020a. We tested associations between categorical variables using Fisher’s exact test. Statistical significance was considered when p values were < 0.05 for single comparisons, or the q-values (Benjamini-Hochberg adjusted p-values) were < 0.05 for multiple comparisons.

## Data Availability

The data that support the findings of this study are available from the following repositories: cBioPortal (https://www.cbioportal.org/), Genomics of Drug Sensitivity in Cancer (https://www.cancerrxgene.org/), and in the Supplementary Files of this article

## List of Abbreviations

TCGA: The Cancer Genome Atlas
GO: Gene Ontology
CB: combined score
KEGG: Kyoto Encyclopaedia of Gene and Genomes
KEA: Kinase Enrichment Analysis
ChEA: Chromatin Immunoprecipitation Enrichment Analysis
X2K: Expression2Kinases
HPV: Human Papillomavirus

## Declarations

### Author Contributions

The study was conceptualised by DK, PN, MZ, and MS; The formal methodology was devised by DK, PN, MZ, EZ, RT, VD, ZN, SM, and MS; DK, PN, RT, VD, ZN, EZ, SM, and MS performed the formal analysis of the datasets; DK, MZ, VD, RT, ZN, EZ, SM, and PN wrote the draft manuscript; the manuscript was revised by DK, PN, MZ, RT, VD, ZN, EZ, SM, and MS; DK, PN, and MS created visualisations.

## Ethics Approval

The study protocol was by the University of Cape Town; Health Sciences Research Ethics Committee IRB00001938 of this study. The analyses in this study utilised publicly available datasets that were collected by the TCGA and GDSC from consenting participants. Here, all analyses were performed following the relevant policies, regulations and guidelines provided by the TCGA and GDSC for analysing their datasets and reporting of the findings.

## Competing interests

The authors declare that they have no competing interests

## Notes

### Competing Interest Statement

The authors have declared no competing interest.

### Funding Statement

Not Applicable

### Author Declarations

The study protocol was by the University of Cape Town; Health Sciences Research Ethics Committee IRB00001938 of this study.

